# Circulating interleukin-6 levels and incident ischemic stroke: a systematic review and meta-analysis of population-based cohort studies

**DOI:** 10.1101/2021.03.27.21254451

**Authors:** Andreas Papadopoulos, Konstantinos Palaiopanos, Harry Björkbacka, Annette Peters, James A. de Lemos, Sudha Seshadri, Martin Dichgans, Marios K. Georgakis

## Abstract

**Objective:** To determine the association between circulating interleukin-6 (IL-6) levels and risk of incident ischemic stroke in the general population.

**Methods:** Following the PRISMA guidelines, we systematically searched the literature for population-based prospective cohort studies exploring the association between circulating IL-6 levels and risk of incident ischemic stroke. We pooled association estimates for ischemic stroke risk with random-effect meta-analyses and explored non-linear effects in dose-response meta-analyses. Risk of bias was assessed with the Newcastle-Ottawa scale (NOS).

**Results:** We identified 11 studies (n=27,411 individuals; 2,669 incident stroke cases) meeting our eligibility criteria. Overall, quality of the included studies was high (median 8 out of 9 NOS points). In meta-analyses, 1-standard deviation increment in circulating IL-6 levels was associated with a 19% increase in risk of incident ischemic stroke over a mean follow-up of 12.4 years (RR 1.19; 95% CI 1.10 to 1.28). A dose-response meta-analysis showed a linear association between circulating IL-6 levels and ischemic stroke risk. There was only moderate heterogeneity and the results were consistent in sensitivity analyses restricted to studies of low risk of bias and studies fully adjusting for demographic and vascular risk factors. The results also remained stable following adjustment for publication bias.

**Conclusions:** Higher circulating IL-6 levels in community-dwelling individuals are associated with higher long-term risk of incident ischemic stroke in a linear pattern and independently of conventional vascular risk factors. Along with findings from genetic studies and clinical trials, these results provide additional support for a key role of IL-6 signaling in ischemic stroke.

## INTRODUCTION

Stroke is a leading cause of adult disability and mortality worldwide.^1,2^ Identifying risk factors for stroke is important for developing effective primary and secondary preventive strategies. Inflammation has recently attracted attention as a potential target for lowering ischemic stroke risk.^3,4^ Data from large-scale trials^5–7^ have provided proof-of-concept evidence that anti-inflammatory approaches can lower cardiovascular risk. Still, these trials tested combined cardiovascular endpoints and evidence regarding the utility of anti-inflammatory approaches specifically for stroke prevention is scarce.^8^

Developing effective anti-inflammatory approaches for stroke prevention would require identifying key inflammatory mediators involved in stroke pathogenesis.^9,10^ While there is extensive literature regarding association of C-reactive protein levels, a general marker of inflammation, with stroke,^11^ there is only limited data regarding other inflammatory cytokines. Data from human genetic studies have suggested a potentially causal role of the pro-inflammatory cytokine interleukin-6 (IL-6) in vascular disease,^12–14^ thus making it a promising drug target.

Moving towards anti-inflammatory treatments specifically targeting IL-6 signaling^15^ would benefit from clarifying the magnitude and shape of the association between circulating IL-6 levels and ischemic stroke. While prospective cohort studies have established robust associations between circulating IL-6 levels with risk of coronary artery disease,^16^ there is only limited evidence regarding associations with ischemic stroke,^17–19^ which also entails mechanisms other than atherosclerosis. Here, we set out to leverage data from published literature along with unpublished cohort studies in a systematic review and meta-analysis in order to explore the association of circulating IL-6 levels and risk of incident ischemic stroke in population-based prospective cohort studies.

## METHODS

This systematic review was conducted according to the Preferred Reporting Items for Systematic Reviews and Meta-Analyses (PRISMA) statement guidelines.^20^

### Search strategy

Two independent reviewers (A.P. and K.P.) systematically screened the medical search engine PubMed from inception to March 4, 2021. We searched for cohort studies investigating the association between circulating IL-6 levels and risk of ischemic stroke using a combination of the predefined key words “interleukin-6”, “IL-6”, “stroke”, and “cerebrovascular disease”. Reference lists of eligible articles were hand-searched for possible eligible studies not identified through the primary database search (“snowball” procedure). No language or publication year restrictions were applied. Eligible studies were assessed for potential population overlap according to recruitment period, geographical site, study name, and sample size. In case of overlap, we included the study with the largest number of incident events. We also contacted the corresponding authors of studies, which did not present the desired analysis but presented the required variables, to request for additional data.

### Eligibility criteria

Eligible studies should be of a prospective cohort design. Case-cohort and nested case-control analyses within prospective cohorts, as well as prospective *post hoc* analyses from randomized clinical trials, were also considered eligible. Case-control and cross-sectional studies, case series, case reports, systematic or narrative reviews, as well as animal and *in vitro* studies were excluded. Eligible studies should be preferably based on the general population. Studies on high-risk populations, such as populations with conventional vascular risk factors (e.g. diabetes mellitus or hypertension), but free of stroke at baseline were also included. Studies including solely individuals with a history of stroke were excluded, as did studies in very specific high-risk populations, such as individuals with advanced chronic kidney disease on hemodialysis.

The exposure variable of interest was circulating IL-6 levels quantified in plasma or serum by immunoassay methods. Due to the lack of universally accepted IL-6 normal value range and differences across variable laboratory kits used by individual studies, we analyzed IL-6 in standardized (1-standard deviation (SD) increment) and not absolute values.

We included studies that explored associations between circulating IL-6 levels and risk of incident ischemic stroke, defined according to standardized clinical criteria. We excluded studies examining associations with: (i) a combined cardiovascular endpoint also including ischemic stroke, but not providing association estimates for ischemic stroke; (ii) clinically silent brain infracts; (iii) stroke mortality; (iv) TIAs; (v) recurrent stroke; and (vi) hemorrhagic stroke. Studies examining combined endpoints of ischemic and hemorrhagic stroke or ischemic stroke and TIAs were included on the basis of the fact that the majority of acute cerebrovascular events represent ischemic strokes.^21,22^ Prospective studies examining ischemic stroke events over follow-up, but not excluding individuals with a history of prevalent stroke at baseline, were also included in our review, as long as such individuals represented the minority (<50%) of the baseline population.

### Data extraction

A predefined spreadsheet was used to extract the following variables from each eligible study: publication details (author, year), study parameters (geographical origin, recruitment period, design, sample size, follow-up information), demographic population characteristics (age, sex, race), baseline cardiovascular risk factors (body mass index, diabetes mellitus, atrial fibrillation, coronary artery disease, hypertension, smoking status, hypercholesterolemia), IL-6 quantification details (sample, laboratory kit, storage temperature, scale of qualification), ischemic stroke assessment (definition, clinical scales used, imaging modality, number of cases), and statistical analysis details (analysis type, effect estimates, 95% CI, adjusting variables). Where supplementary data were needed, the corresponding author was contacted.

### Risk of bias assessment

We evaluated studies for risk of bias using the nine-item cohort subscale of the Newcastle-Ottawa scale.^23^ The following criteria were assessed: (i) representativeness of the exposed cohort: a point was awarded when individuals were drawn from the general population and included both males and females; (ii) selection of the non-exposed cohort: a point was awarded when individuals were drawn from the same community as the exposed; (iii) exposure ascertainment: a point was awarded when the blood drawing protocol and the kit used for IL-6 quantification were reported; (iv) outcome presence at start of study: a point was awarded when individuals with a history of ischemic stroke at baseline were excluded from the analysis; (v and vi) two items for comparability: one point was awarded if the study adjusted for age and sex, and a second point was awarded if the study additionally adjusted for conventional vascular risk factors (at least lipids, blood pressure, diabetes, and body mass index); (vii) outcome assessment: a point was awarded when the study presented association estimates specifically for ischemic stroke, excluding TIAs and hemorrhagic strokes, as well as when a clear definition was provided and each event was confirmed by at least a trained physician; (viii) length of follow-up: a point was awarded when the mean or median follow-up of the cohort was >5 years; (ix) adequacy of follow up cohorts: a point was awarded when the attrition rate was lower than <10%.

### Statistical analysis

For each eligible study we extracted association estimates and 95%CI between circulating IL-6 levels and incident ischemic stroke. Out of the 11 studies included in our meta-analysis, 8 presented hazard ratios (HR), 2 odds ratios (OR) and 1 relative risks (RR). First, we transformed all estimates and their corresponding 95% CIs to RRs. If ischemic stroke incidence in the examined cohort exceeded 10%, we used validated formulae,^24^ whereas in studies with <10% incidence, we considered HRs and ORs to be very close to RRs and thus applied no transformation.^24,25^

Seven out of the 11 studies analyzed 1-SD increment in log-transformed IL-6 levels, whereas the remaining 4 studies presented association estimates across tertiles or quartiles of IL-6. To enable a meta-analysis across all studies, we used the method of generalized least squares for trend estimation of summarized dose-response data to derive association estimates per 1-SD increment in studies presenting analyses in tertiles or quartiles.^26,27^ Doses across each category were calculated as fitting SD-increases by using the median values of each tertile/quartile projected on a normal distribution.

We then performed random-effect meta-analyses of the derived association estimates using the method described by DerSimonian and Laird^28^ and obtained a pooled RR with 95%CI for the risk of incident ischemic stroke per 1-SD increase in IL-6 levels. The presence of heterogeneity was evaluated by the I^2^, calculated via the Cochran Q statistic. We defined low, moderate, and high heterogeneity as an I^2^ of <25%, 25% to 75%, and >75%, respectively.^29^

To explore the robustness of our findings, we carried out sensitivity analyses restricted to: (i) studies of the general population; (ii) studies not including TIAs as an outcome; (iii) studies exclusively exploring ischemic stroke: (iv) studies excluding individuals with a history of prevalent stroke at baseline; (v) studies providing imaging confirmation (via CT or MRI) for an infarction beyond the clinical definition of ischemic stroke; (vi) studies adjusting their results for demographic and conventional vascular risk factors; (vii) studies additionally adjusting for circulating C-reactive protein levels; and (viii) studies of a follow-up ≥5 years; (ix) studies fulfilling at least 8 out of the 9 quality criteria of NOS. We also ran a separate analysis of the seven studies using the High Sensitivity 600 Quantikine ELISA kit by R&D Systems for quantifying circulating IL-6 levels to avoid heterogeneity in the effects due to differences in the used kit. Finally, to exclude potential outlier effects of individual studies, a “leave-one-out” sensitivity analysis was performed.

Furthermore, we sought to examine whether circulating IL-6 levels follow a linear association with the risk of incident ischemic stroke. We used multivariate random-effect meta-analysis and constructed a double-tail restricted three-cubic knot (10%, 50%, 90%) flexible model, which demonstrates the actual shape of the relationship between the RR for incident ischemic stroke plotted against IL-6 percentiles.^30^

The effect of potential publication bias (small-study effects) was explored using the Egger’s test.^31^ In cases of evidence of small-study effects (P>0.10), we further adjusted the pooled effect estimate for publication bias using a “trim and fill” analysis.^32^ The results were graphically presented with a funnel plot.

All analyses were conducted with the STATA Software, version 16.1 (Stata Corporation, College Station, TX, USA).

### Data availability

Data not provided in the article due to space limitations will be made available upon reasonable request to the corresponding author.

## RESULTS

### Review of literature

**Figure 1** summarizes the study selection process. Following an initial screening of 2,702 articles yielded by the literature search, we identified 8 articles,^17–19,33–36^ referring to 11 individual studies (n=27,411), meeting our eligibility criteria. Four of the included studies (DHS, FHS-offspring, MONICA/KORA and MDCS-CV) have not published results on the associations between circulating IL-6 levels and risk of ischemic stroke, but the respective data were provided as part of secondary analysis in a recent meta-analysis focusing on the association of monocyte chemoattractant protein-1 and stroke.^37^ Association estimates from the latter meta-analysis were used for the purposes of the current study.

**Figure 1.**
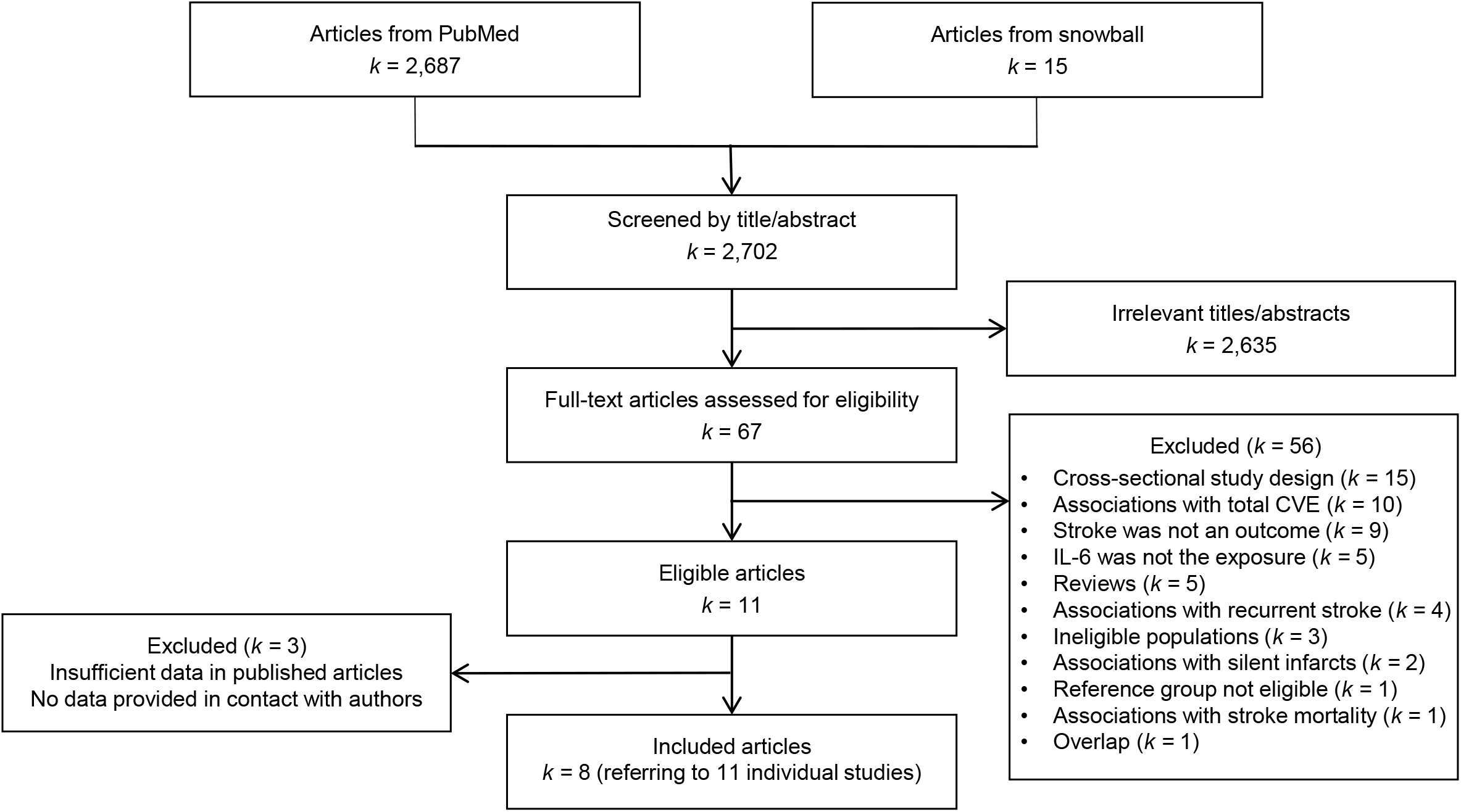
Flowchart of the study selection process. Steps and number of articles screened per step during the study selection process. CVE = cardio-vascular events.

### Descriptive study characteristics and risk of bias assessment

Summarized descriptive characteristics of the 11 included studies are presented in **Table 1** and **in Supplementary Table 1**. Mean age of all individuals was 60.5 years (study range, 44.0 to 75.9 years) and 54.8% of the study participants were females. Mean duration of follow-up was 12.4 years (study range, 3.2 to 20.0). All included studies followed a prospective study design: seven of them (n=21,384) featured a cohort study design, while the remaining 4 studies presented either case-cohort (k=2, n=3,425) or nested case-control (k=2, n=2,602) analyses within larger cohorts. Nine of the studies (n=26,149) were based on general population individuals, while 2 (n=1,262; OSAKA and PROSPER) were restricted to high-risk individuals with at least one established vascular risk factor. IL-6 measurements were made on blood drawn at baseline (stored at −70 to −80° C until analyzed). Four studies (n=7,813) used serum and 4 (n=10,813) used plasma samples to quantify IL-6, whereas the exact sample was not reported in 3 studies. The kit most commonly used for IL-6 measurements was the High Sensitivity 600 Quantikine ELISA by R&D Systems (7 studies; n=16,918). In 9 studies, where this was reported, intra- and inter-assay coefficients of variation were ≤10%.

**Table 1.**
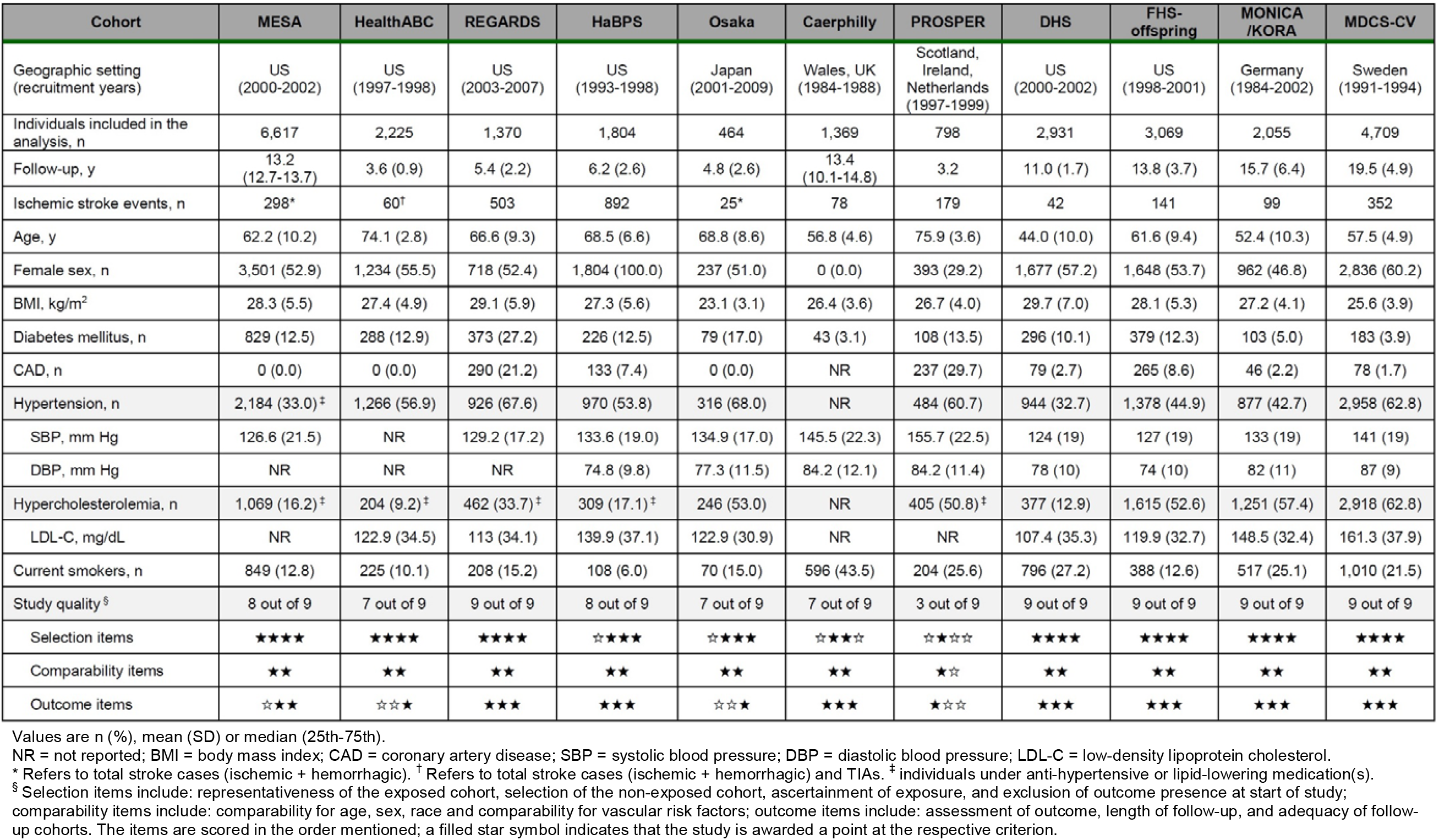
Descriptive characteristics of the included studies. Characteristics of studies investigating the association between circulating blood IL-6 values and prospective stroke.

Regarding outcome assessment, 9 studies (n=25,244) excluded patients with a history of stroke at baseline and 8 studies (n=18,105) specifically addressed ischemic stroke as their outcome excluding patients with hemorrhagic stroke or TIA. All endpoints were validated by at least two trained physicians, who reviewed each patient’s medical or autopsy files. Only 4 studies (n=4,436) explicitly required imaging confirmation of an infarction with CT and/or MRI, as definition of ischemic stroke.

The overall study quality was high, with 5 of the studies (45.5%) fulfilling all 9 criteria of the Newcastle-Ottawa scale criteria a (**Table 1**). The median quality score was 8 out of 9 (IQR 2, range 3 to 9). The items “representativeness of the exposed cohort”, “outcome not present at start of study”, “assessment of outcome” and “length of follow-up” accounted for most non-awarded points. All studies controlled for age, sex (if applicable) and race (if applicable) and all but one of the studies additionally controlled for conventional vascular risk factors.

### Circulating IL-6 and risk of incident ischemic stroke

In the meta-analysis of the 11 studies, we found a 1-SD increment in circulating IL-6 levels at baseline to be associated with a 19% higher risk of incident ischemic stroke over a mean follow-up of 12.4 years (RR 1.19; 95% CI 1.10 to 1.28; 27,411 individuals; 2,669 events, **Figure 2**). The results remained stable in sensitivity analyses for studies excluding individuals with history of prevalent stroke at baseline, studies focusing on incident ischemic stroke explicitly excluding cases of TIA and hemorrhagic stroke, as well as studies requiring imaging confirmation of an infarction (**Figure 3**). Furthermore, our analyses revealed that controlling for conventional vascular risk factors yielded the same pooled association estimate as our main analysis. Further adjustment for high-sensitivity CRP levels led to an anticipated attenuation of the association estimate, as CRP is downstream of IL-6,^15^ but the association remained statistically significant. Similar results were also obtained in a sensitivity analysis restricted to studies of low risk of bias (scoring at least 8 out of 9 in NOS). Restricting our analyses to studies quantifying IL-6 with the most commonly used High Sensitivity 600 Quantikine ELISA kit did not change the result. Of note, pooling a set of 6 studies of stroke-free individuals in the general population that specifically examined over a follow-up of ≥5 with years associations with incident ischemic stroke (excluding TIAs and hemorrhagic strokes) and further adjusted for conventional vascular risk factors on top of age, sex and race yielded similar results (RR 1.16; 95% CI 1.07 to 1.25; 6 studies; 15,938 individuals; 2,029 events). Finally, in “leave-one-out” sensitivity analyses, we found no evidence that any single study significantly influenced the results of our main analysis (**Supplementary Figure 1**).

**Figure 2.**
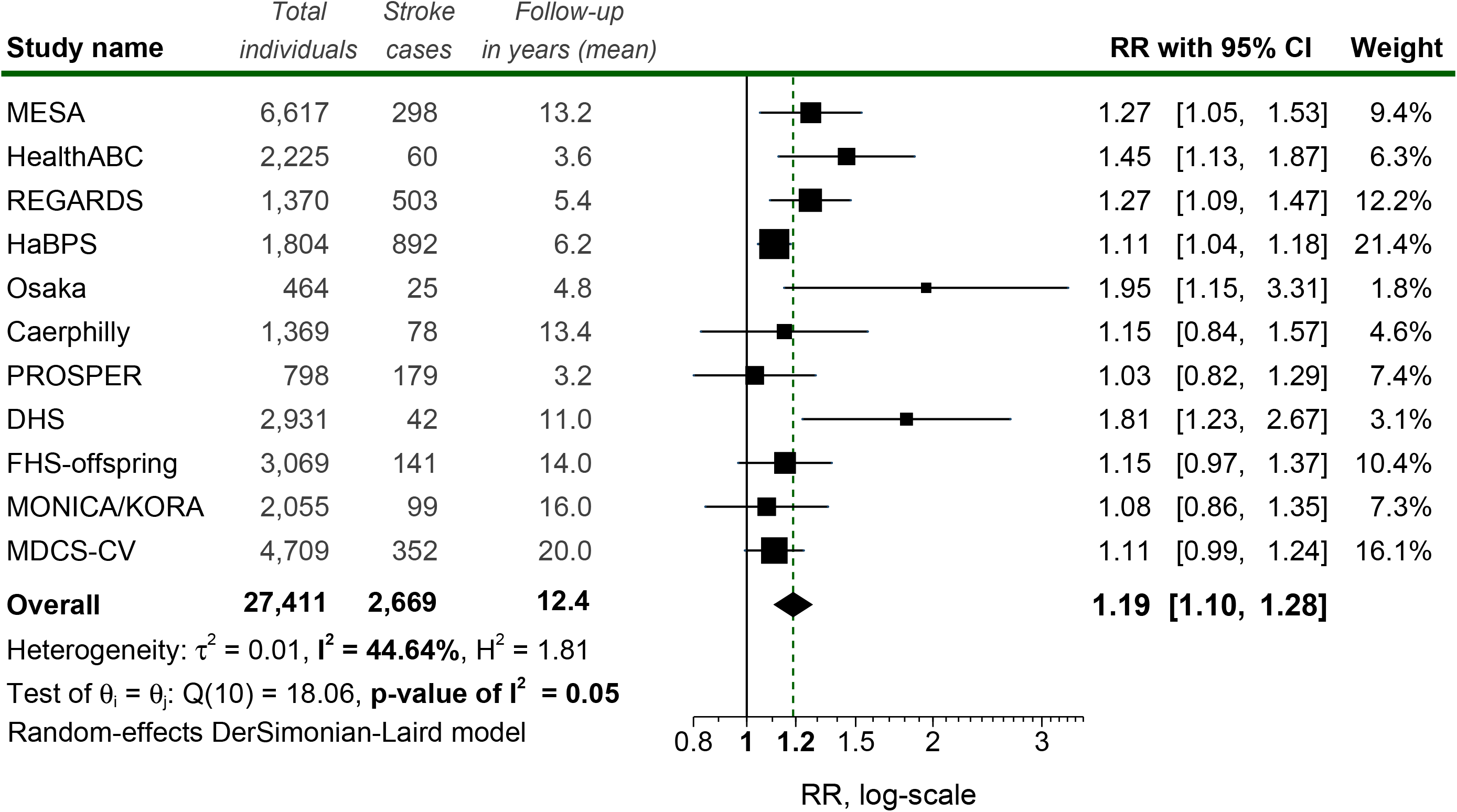
Meta-analysis of the association between circulating IL-6 levels (1-SD increment) and risk of incident ischemic stroke. Risk Ratios (RR) of each study are depicted as data markers; black boxes around the data markers indicate the statistical weight of the respective study; 95% CIs are indicated by the black error bars; pooled-effect estimate along with its 95% CI is reflected as a black diamond.

**Figure 3.**
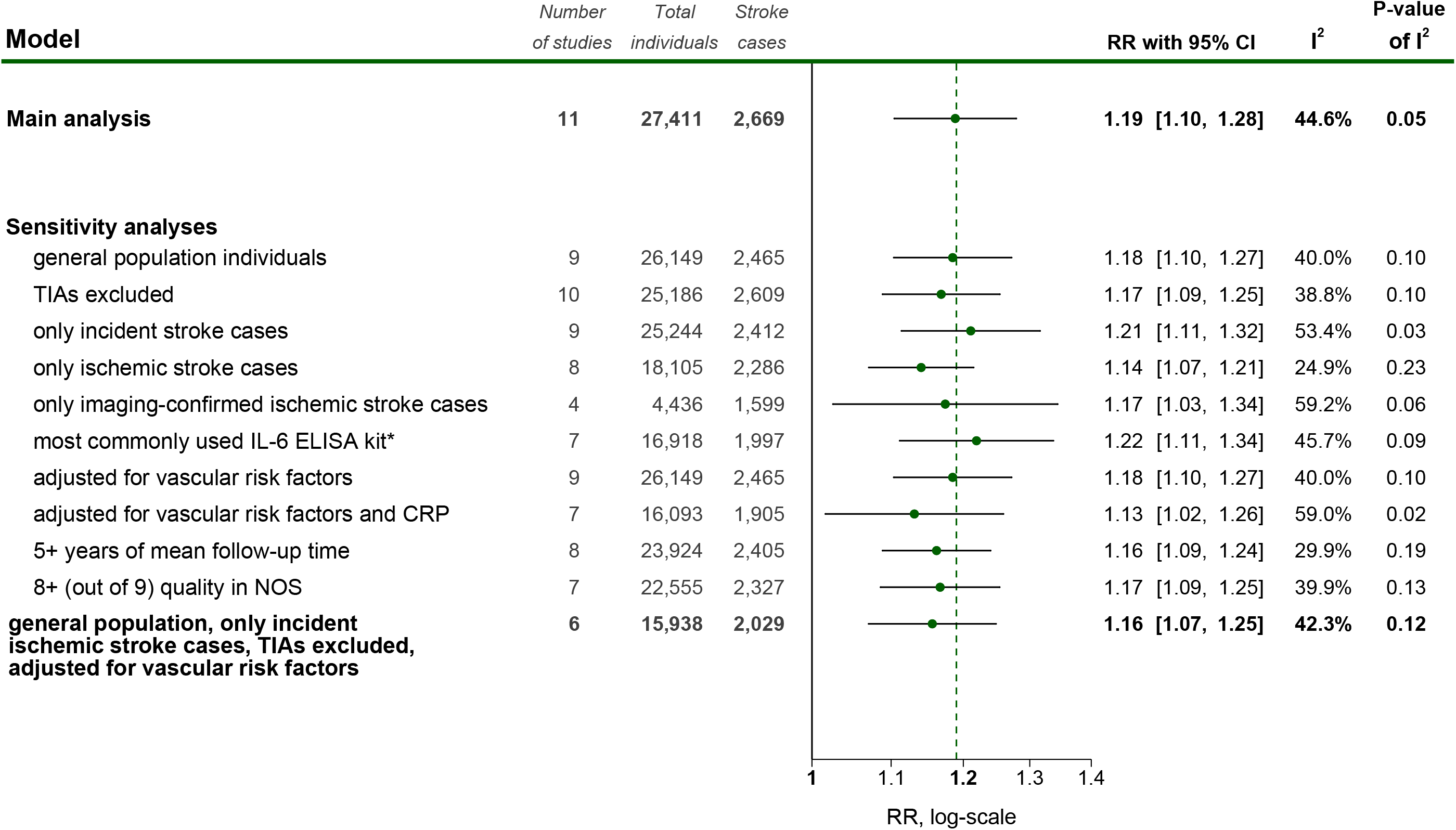
Sensitivity analyses of the association between circulating IL-6 levels (1-SD increment) and risk of incident ischemic stroke. Pooled random-effect Risk Ratios (RR) of each analysis are presented as green data markers; 95% CIs are indicated by the black error bars; the vertical green dashed line indicates the overall effect estimate of the main analysis. * High Sensitivity 600 Quantikine ELISA by R&D Systems. ELISA = enzyme-linked immunosorbent assay; CRP, C-reactive protein; NOS = Newcastle - Ottawa scale.

There was only moderate heterogeneity in the main analysis (I^2^=44.6%, p=0.05; **Figure 2**), which was not entirely resolved in any of the sensitivity analyses (**Figure 3**). The funnel plot for our main analysis is presented in **Supplementary Figure 2**. Although the Egger’s test detected small-study effects (P=0.03) indicating potential presence of publication bias, the association between circulating IL-6 levels and incident ischemic stroke remained stable (RR 1.14; 95% CI 1.05 to 1.24) after correcting our analysis for small-study effects with the “trim and fill” method.

As a final step, we aimed to explore the shape of the association between circulating IL-6 levels and risk of incident ischemic stroke. In a dose-response meta-analysis including data from 5 studies (13,385 individuals; 1,831 events), we found a linear relationship between circulating IL-6 levels and incident ischemic stroke (p for non-linearity: 0.52; **Figure 4**).

**Figure 4.**
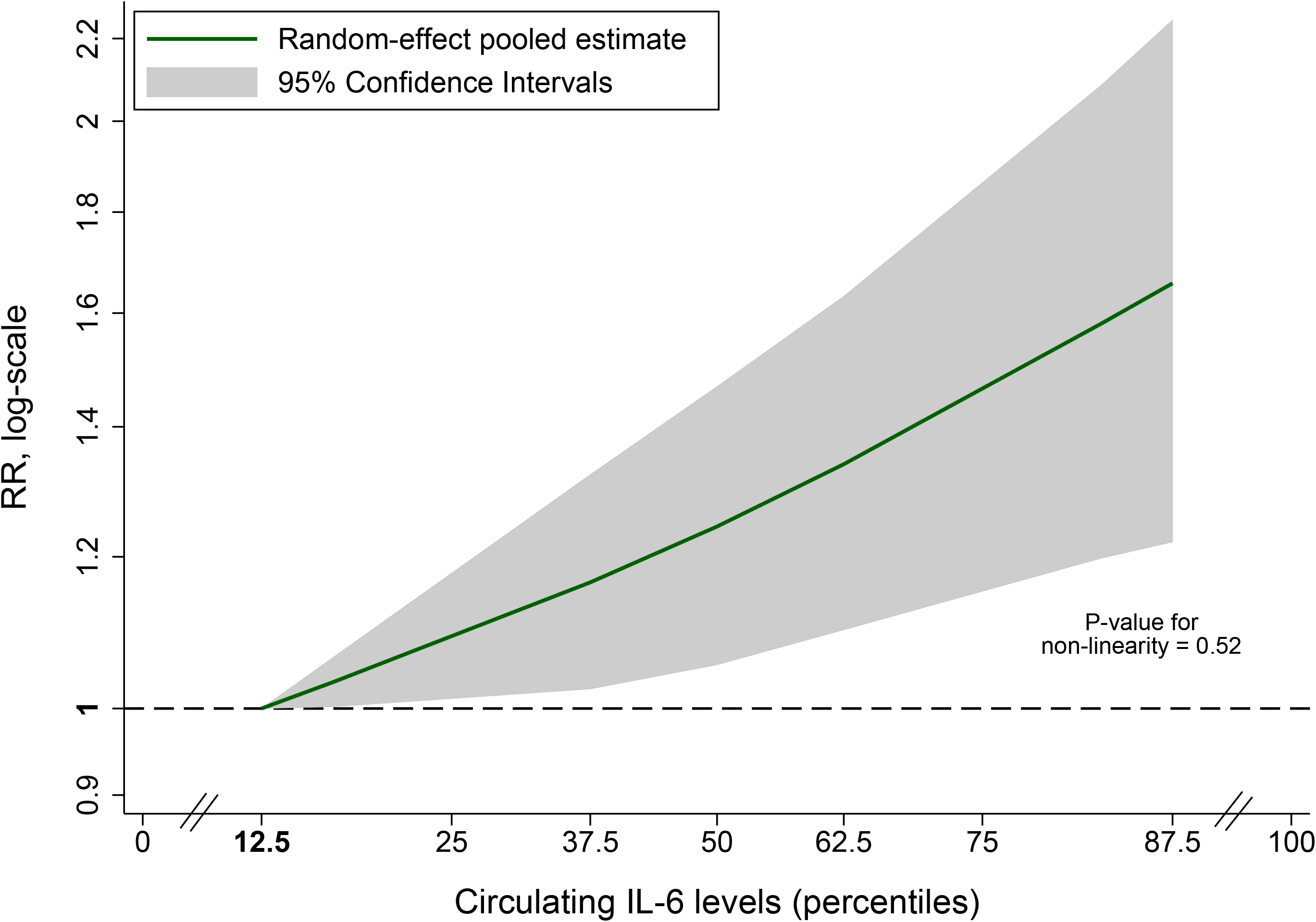
Dose-response meta-analysis of the association between circulating IL-6 levels (standardized values in percentiles) and risk of incident ischemic stroke. A double-tail restricted, 3 cubic knot (10%, 50%, 90%) flexible model was used. IL-6 values have been projected on a normal distribution and are presented as percentiles. The median of the 1^st^ quartile (12.5^th^ percentile) is used as the reference. The analysis is based on 5 studies (13,385 individuals; 1,831 stroke cases).

## DISCUSSION

Pooling data from 11 population-based prospective cohort studies involving 27,411 individuals and 2,669 stroke events, we found higher circulating IL-6 levels at baseline to be associated with a higher risk of incident ischemic stroke over a mean follow-up of 12.4 years. IL-6 levels showed a linear relationship with the risk of ischemic stroke following a dose-response pattern. Overall, the study quality was high and the results were stable in sensitivity analyses, as well as when correcting for publication bias.

Our meta-analysis extends previous data related to the associations between circulating IL-6 levels with acute coronary events and other vascular phenotypes^16^ to ischemic stroke. IL-6 signaling has been demonstrated as one of the most promising targets for anti-inflammatory approaches in cardiovascular disease. The CANTOS trial tested canakinumab, a monoclonal antibody against IL-1b, which is upstream to IL-6, in patients with a recent myocardial infarction and showed a beneficial effects against a combined cardiovascular endpoint, also involving stroke.^5^ Interestingly, secondary analyses from CANTOS showed that the benefit was restricted to individuals in whom canakinumab resulted in meaningful reductions in IL-6 levels.^38^ Still, CANTOS could not specifically show benefit against stroke,^5^ possibly as a result of limited power. The results from the current meta-analysis, when seen together with Mendelian randomization results supporting associations between lifetime genetically downregulated IL-6 signaling and lower ischemic stroke risk^13^ provide further support in favor of IL-6 signaling as a promising target for lowering stroke risk.

An interesting finding from our analysis is the clearly log-linear dose-response relationship between IL-6 levels and stroke risk. Our results indicate an approximately 19% increment in risk of ischemic stroke per-SD increment in log-IL-6 levels. This magnitude of effect, along with the clear dose-response pattern, is comparable to the magnitude and shape of associations that have been reported for non-HDL cholesterol levels (12%, 95%CI: 4-20%),^39^ and systolic blood pressure (24%, 95%CI: 15-35%),^40^ both key therapeutic targets for lowering ischemic stroke risk. While this association estimate slightly reduced after correcting for publication bias, it still remained in the same order of magnitude (RR 1.14), thus supporting a meaningful association with the risk of ischemic stroke.

Our results should be viewed in the context of specific methodological strengths. First, this meta-analysis is clearly based on prospective cohort population-based studies with relatively long follow-up period, thus precluding the possibility of reverse causation. Furthermore, our rigorous search of published literature, allowed us to pool a large sample size including more than 2,600 incident stroke cases, thus offering the power to explore interesting aspects of this association, such as its robustness against specific forms of bias in sensitivity analyses, and dose-response relationships. Finally, as observed in our risk of bias analysis, the quality of the included studies was generally high, thus further supporting to the validity of the results.

Our study also has limitations. There was moderate heterogeneity in the main analysis (*I*^*2*^=45%), which points to key methodological differences between individual studies. Specifically, while all of the studies had a prospective study design, some of them applied case-cohort or nested case-control approaches within the larger cohorts. Furthermore, there were wide differences with regards to mean follow-up intervals ranging from 3 to 20 years across studies. Similarly, there were between-study differences regarding the definition of the outcomes, with some of the studies focusing only on stroke as a whole and not ischemic stroke, whereas other studies also included transient ischemic attacks. Still, it should be mentioned that the results were stable in all sensitivity analyses, even the one focusing on imaging-confirmed infarctions. Another source of heterogeneity is the method used for measuring IL-6 levels, for which, as opposed to high-sensitivity CRP, there are no standard clinical platforms for quantification. To address this issue, we performed all our analyses based on standardized IL-6 levels, but still differences between studies might persist and might affect the results, especially those from the dose-response meta-analysis. Finally, there was evidence of small-study effects indicating publication bias in our analysis, but the results were stable after correcting for it with the trim and fill method.

In summary, as illustrated in our meta-analyses, data from observational studies support a clear dose-response association between circulating IL-6 levels and risk of incident ischemic stroke among stroke-free individuals at baseline. While these results cannot establish causality, when triangulated with evidence from human genetic data, as well as indirect evidence from clinical trials, they provide further support for IL-6 signaling as a promising target for lowering the risk of ischemic stroke.

## Supporting information

PRISMA checklist

## Appendix 1: Authors

| **Name** | **Location** | **Contribution** |
| --- | --- | --- |
| Andreas Papadopoulos, MD | General Military Hospital of Athens, Greece | data acquisition, analysis, and interpretation of data; statistical analysis; drafting of the manuscript |
| Konstantinos Palaiopanos, MD | National Public Health Organization, Athens, Greece | data acquisition, analysis, and interpretation of data; drafting of the manuscript |
| Harry Björkbacka, PhD | Lund University, Malmö, Sweden | data acquisition; critical revision of the manuscript for intellectual content; study supervision |
| Annette Peters, PhD | Helmholtz Zentrum München, Germany | data acquisition; critical revision of the manuscript for intellectual content; study supervision |
| James A. de Lemos, MD | University of Texas Southwestern Medical Center, Dallas, TX, USA | data acquisition; critical revision of the manuscript for intellectual content; study supervision |
| Sudha Seshadri, MD | University of Texas Health Sciences Center, San Antonio, TX, USA | data acquisition; critical revision of the manuscript for intellectual content; study supervision |
| Martin Dichgans, MD | LMU Munich, Germany | interpretation of data; critical revision of the manuscript for intellectual content; study supervision |
| Marios K. Georgakis, MD, PhD | LMU Munich, Germany | concept and design; data acquisition, analysis, and interpretation of data; drafting of the manuscript; critical revision of the manuscript for intellectual content; study supervision |

