## Supplementary Materials for "Circulating interleukin-6 levels and incident ischemic stroke: a systematic review and meta-analysis of population-based cohort studies"

| **Cohort** | **Study design** | **Population characteristics** | **IL-6 measurement** | **Stroke assessment and definition** | **Analysis presented** |
| --- | --- | --- | --- | --- | --- |
| Multi-Ethnic Study of Atherosclerosis (MESA) | Cohort | Multi-ethnic (non-Hispanic Whites, African Americans, Hispanics, Chinese Americans); general population; US (New York, Baltimore, Chicago, Los Angeles, Twin Cities, Winston-Salem); free of clinical CVD at baseline | HS600 kit* in fasting blood samples (stored at -80°C) with analytical CV=6.3% | All stroke events; only incident; TIAs excluded. Individuals were regularly contacted and have been seen in person at 4 examinations. Death certificates and autopsy reports were reviewed. Outpatient summaries/diagnostic test results were also reviewed. Three vascular neurologists examined all events possibly related to TIA or stroke. Stroke was defined as rapid onset of neurologic deficit, headache, or meningism AND Neurologic deficits not secondary to brain trauma, tumor, infection, or other non-vascular cause AND Clinically relevant lesion on brain imaging OR Duration >24h OR Death at ≤24h | Cox proportional hazards; HRs for T2, T3 Vs T1 |
| Health, Aging, and Body Composition (HealthABC) study | Cohort | General population; US (Pittsburgh, Memphis); free of clinical CVD at baseline | HS600 kit* in fasting serum samples (stored at -70°C) with inter-assay CV=10.3% | All stroke events; only incident. Individuals were regularly contacted and seen in person annually. When an event was reported, hospital records were examined by an adjudicator at each site. Stroke was defined as any overnight hospitalization for definite or probable stroke or death with stroke as underlying cause. TIAs were not explicitly excluded. | Cox proportional hazards; RR per 1-SD increase in log-transformed IL-6 and RRs for T2, T3 Vs T1 |
| Reasons for Geographic and Racial Differences in Stroke (REGARDS) study | Case-cohort; cohort random sample, was weighted to analytic cohort (~3% stroke incidence was present) | Two-ethnic (blacks - oversampled: 42%, whites); general population; all over the US; oversampled: 56% the stroke buckle (coastal plains of North Carolina, South Carolina, Georgia) and the stroke belt (rest of North, South Carolina, Georgia, Tennessee, Mississippi, Alabama, Louisiana, Arkansas); stroke-free at baseline | HS600 kit* in fasting blood samples (stored at -80°C) with analytical CV=6.3%. | Only ischemic stroke; only incident; TIAs excluded; only imaging-confirmed ischemic stroke cases. Individuals were regularly contacted. Medical records were obtained for death or suspicion of stroke or TIA. After pre-review by a trained stroke clinician, a committee of stroke physicians reviewed records to validate strokes. Strokes were defined as focal neurologic symptoms lasting >24h or non-focal symptoms with positive imaging for stroke. Stroke events were classified ischemic or hemorrhagic and secondarily by ischemic subtypes. Hemorrhagic strokes were excluded. | Weighted (according to the inverse sampling probability in the cohort random sample) Cox proportional hazards; HRs for Q2, Q3, Q4 Vs Q1 |
| Hormones and Biomarkers Predicting Stroke (HaBPS) study | Nested case-control; controls matched for age, race, date of enrollment, follow-up time; individuals drawn from the Women's Health Initiative study | Female general population; based around 40 centers throughout the US; stroke-free at baseline | HS600 kit* in fasting plasma samples (stored at -70°C) with analytical CV=7.6%. | Only ischemic stroke; only incident; TIAs excluded; only imaging-confirmed ischemic stroke cases. Individuals were contacted annually. Medical records were collected for potential stroke. Adjudication was performed locally (trained physicians) and centrally (study neurologists). Ischemic stroke was defined as the rapid onset of a persistent neurologic deficit attributed to an obstruction lasting >24h and without evidence for other causes. Only stroke events requiring hospitalization were considered. Stroke events were classified as ischemic or hemorrhagic on review of reports of brain imaging studies. | Multivariate unconditional logistic regression; ORs for Q2, Q3, Q4 Vs Q1 |
| Osaka Follow-up Study for Carotid Atherosclerosis 2 (OSACA2) study | Cohort | Japan (Osaka); ≥1 established cardiovascular risk factor at baseline; free of clinical CVD at baseline | HS600 kit* in serum samples (stored at -80°C) with intra- and inter-assay CV of 7.8% & 7.2%, respectively | Only ischemic stroke; only incident; TIAs excluded; only imaging-confirmed ischemic stroke cases. Individuals were contacted and/or seen in person regularly. Medical records were reviewed independently by 3 physicians if a cardiovascular event was reported. Stroke was defined as an acute disturbance of focal neurological dysfunction with symptoms lasting >24h (or death at ≤24h), including surgical / endovascular treatment resulting from TIAs. TIAs were otherwise excluded. Ischemic and hemorrhagic strokes were confirmed by neuroimaging. Hemorrhagic strokes were excluded. | Cox proportional hazards with stepwise multivariable regression (backward elimination); HR per 1-SD increase in log-transformed IL-6 |
| Caerphilly Study | Cohort | Male general population; UK, Wales (Caerphilly and Speedwell) | HS600 kit* in fasting plasma samples (stored at -70°C) with intra- and inter-assay CV 7% & 8%, respectively | Only ischemic stroke; TIAs excluded. Individuals were flagged with the National Health Service Central Registry. ICD-9 codes: 430-438 defined fatal stroke. ICD-9 codes: 430–436 were searched on hospital records to identify non-fatal strokes. Ischemic stroke events were validated from 2 (3 if disagreement) stroke clinicians. Hemorrhagic strokes were excluded. | Cox proportional hazards with strata defined by presence / absence of CVD at baseline; HRs for T2, T3 Vs T1 |
| Prospective Study of Pravastatinin the Elderly (PROSPER) | Nested case-control; controls matched for age, gender, treatment allocation | Scotland (Glasgow), Ireland (Cork), Netherlands (Leiden); half with ≥1 established cardiovascular risk factor at baseline, half with established clinical CVD at baseline | Laboratory measurements in fasting blood samples | Only ischemic stroke; TIAs excluded; only imaging-confirmed ischemic stroke cases. Individuals were seen in person regularly. Ischemic stroke was defined as one of the following: (1) Rapid onset of focal neurologic deficit lasting >24h or leading to death with neuroimaging showing infarction or no abnormality, or autopsy findings showing infarction, (2) Rapid onset of global neurologic deficit lasting >24h or leading to death with relevant evidence from neuroimaging or autopsy, (3) Focal neurologic deficit (mode of onset uncertain) lasting >24h or leading to death with relevant evidence from neuroimaging or autopsy. | Univariate conditional logistic regression; OR per 1-SD increase in log-transformed IL-6 |
| Dallas Heart Study (DHS) | Cohort; sub-sample of the DHS cohort | Multi-ethnic; general population; US (Dallas County), chosen via a stratified random sample; stroke-free at baseline | In-house IL-6 assay in fasting plasma samples (stored at -80°C) | Only ischemic stroke; only incident; TIAs excluded. Non-fatal stroke was defined by either annual follow-up medical records assessments or tracking hospital admissions through the Dallas–Fort Worth Hospital Council Data Initiative database (coverage 90% of the study region), using ICD-9 codes: 430-438. Fatal stroke was defined by death certification using the National Death Index, using ICD-10 codes: I60-I69. | Cox proportional hazards; HR per 1-SD increase in log-transformed IL-6 |
| Framingham Heart Study (FHS) - Offspring | Cohort; individuals of examination 7 (1998-01) of the FHS-offspring cohort | General population; offspring of the participants of the Original FHS Cohort and their spouses; stroke-free at baseline | HS600 kit* in fasting serum samples (stored at -70°C) with intra-assay CV=3.1% | Only ischemic stroke; only incident; TIAs excluded. Strokes were identified via ongoing clinic and hospital surveillance including medical record review, laboratory testing, imaging, autopsy findings and collaboration with general practitioners, emergency departments and imaging facilities in the area. Ischemic stroke was defined as a focal neurologic deficit lasting >24h with imaging showing an ischemic infarction or no hemorrhage, or if an ischemic infarction was documented at autopsy. | Cox proportional hazards; HR per 1-SD increase in log-transformed IL-6 |
| Monitoring of Trends and Determinants in Cardiovascular Disease sub-cohort of the Cooperative Health Research in the Region of Augsburg (MONICA/KORA) | Case-cohort; cohort random sample was a representative sub-sample of the overall cohort | General population; Germany (Augsburg and surrounding counties); stroke-free at baseline | Sandwich ELISA (CLB, Amsterdam, Netherlands) in serum samples (stored at -80°C) with intra- and inter-assay CV <10% | Only ischemic stroke; only incident; TIAs excluded. Non-fatal stroke was derived by self-report validated by cross-linkage with hospital records and information gathered from the treating physicians. Fatal stroke was defined by death certification using ICD-9 codes: 430-434 (German modified version). | Cox proportional hazards; HR per 1-SD increase in log-transformed IL-6 |
| Malmö Diet and Cancer Study - Cardiovascular (MDCS-CV) | Cohort; a random 50% sub-sample of the MDCS cohort | General population; Sweden (Malmö); stroke-free at baseline | Laboratory measurements in fasting blood samples (stored at -80°C) | Only ischemic stroke; only incident; TIAs excluded. Non-fatal and fatal strokes were defined by record linkage with the National Inpatient Register, the Swedish Causes of Death Register, and the Stroke Register of Malmö (STROMA), according to the ICD-9 codes: 430-438. Ischemic stroke was defined as rapidly developing clinical signs of local or global loss of cerebral functioning lasting >24 h (or death at ≤24h) with neuroimaging verifying the infarction and/or excluding hemorrhage, or if an ischemic infarction was documented at autopsy. | Cox proportional hazards; HR per 1-SD increase in log-transformed IL-6 |

**Supplementary Table 1.** Summary of the study design, population characteristics, methods used for quantifying IL-6 levels, stroke assessment and definitions and analyses presented.

US = United States; CV = coefficient of variation; h = hours; HR = hazard ratio; T = tertile; Q = quartile; CVD = cardiovascular disease; RR = risk ratio; OR = odds ratio; IL-6 = interleukin-6.

*Human IL-6 Quantikine High Sensitivity (HS) ELISA Kit (HS600) by R&D Systems.

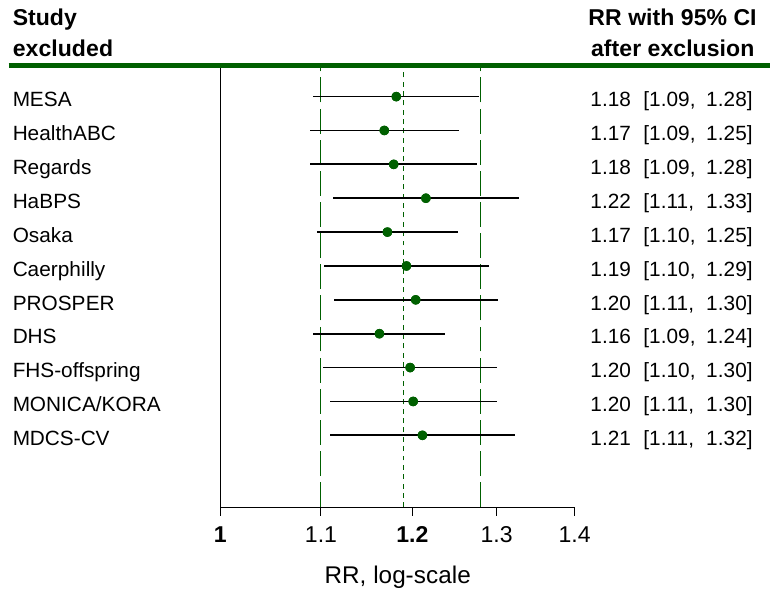
**Supplementary Figure 1.** Leave-one-out sensitivity analyses exploring the effect of each individual study on the overall effect estimate. Green data markers and their corresponding 95% CI error bars indicate the pooled overall effect estimate after excluding the respective study from the analysis.

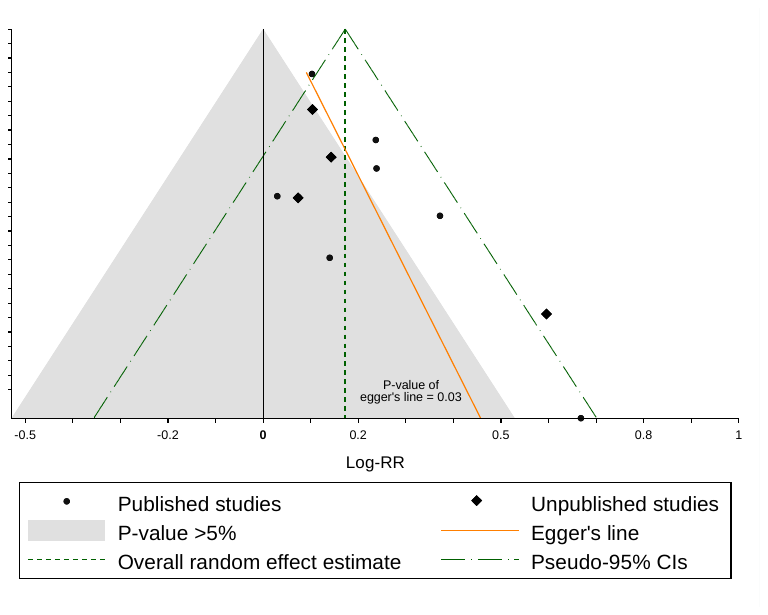

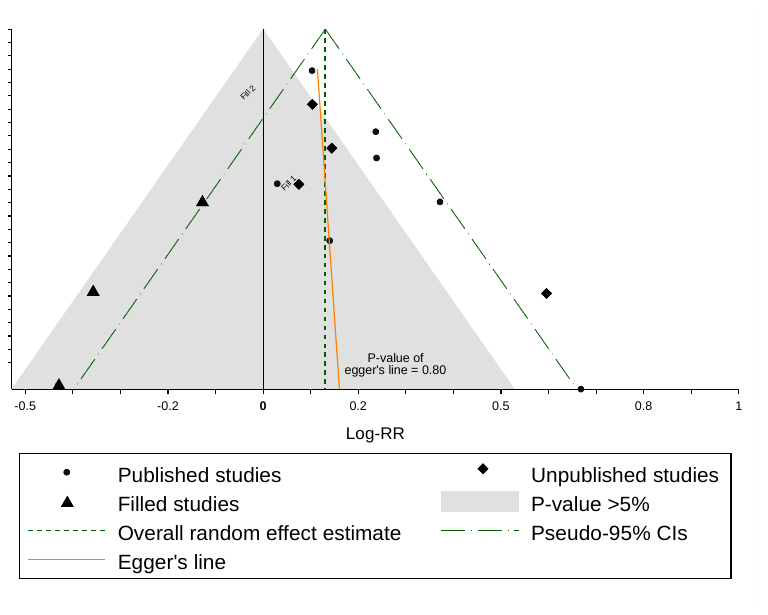

**A.**

**B.**

**Supplementary Figure 2.** Funnel plots of the main analysis. (A) Funnel plot of the 11 studies pooled in the main analysis. Each study is depicted either as a dot (published) or as a diamond (unpublished). The light grey shaded area contains studies with non-significant results at the level of two-sided P=0.05. (B) Funnel plot after adjusting the main analysis with a “trim and fill” approach. Three additional studies (triangles) have been added to account for the small-study effect present in (A).

RR= risk ratio.
